# Use of Computational Phenotypes for Predicting Genetic Subgroups of Cerebral Palsy

**DOI:** 10.1101/2025.02.12.25322169

**Authors:** Imen Alkuraya, Alexandra Santana Almansa, Azubuike Eleonu, Paul Avillach, Annapurna Poduri, Siddharth Srivastava

## Abstract

**Introduction:** Emerging evidence suggests that 20–30% of cases of cerebral palsy (CP) may have a genetic cause. Our group previously identified subsets of patients with CP or CP-masquerading conditions who warrant genetic testing, including those with regression or progressive neurological symptoms (CP masqueraders) and those without any known risk factors for CP (cryptogenic CP). Recognition of these subgroups in clinical settings remains challenging.

**Methods:** To address this challenge, we developed and evaluated a computational phenotyping approach using ICD- 9/ICD-10 billing codes to automatically identify patients with unexplained CP or CP-masquerading conditions who may benefit from genetic testing. We applied this computational phenotyping approach to a cohort of 250 participants from the Boston Children’s Hospital CP Sequencing Study, aimed at identifying genetic causes in CP and CP-masquerading conditions.

**Results:** Manual review served as the gold standard, identifying 8% as CP masqueraders, 42% as cryptogenic CP, and 50% as non-cryptogenic CP. Computational phenotyping based on ICD-9/10 codes achieved a sensitivity of 95%, specificity of 72%, positive predictive value of 77%, and negative predictive value of 94% in identifying cases warranting genetic testing.

**Conclusions:** Our findings demonstrate the feasibility of using computational phenotyping to identify patients with CP or CP- masquerading conditions who warrant genetic testing. Further studies are needed to evaluate the effectiveness and real-world application of this tool in larger healthcare systems. Nonetheless, the computational phenotyping approach holds promise as a possible clinical decision support that could be integrated into electronic health record systems, enhancing clinical workflows and facilitating actionable genetic diagnoses.

## Introduction

Cerebral palsy (CP) is the most common cause of childhood-onset motor disability, affecting 2-3:1000 children in the United States [1,2]. CP is a descriptive term, with multiple possible etiologies [3]. Examples of acquired risk factors for CP include prematurity [4–6], intraventricular hemorrhage [7,8], perinatal stroke [9,10], and hypoxic ischemic encephalopathy [11–13]. There is increasing recognition that genetic factors play a role in the development of CP. Meta-analyses have shown that approximately 20-30% of individuals of CP may have an underlying genetic disorder [14] [15]. However, due to past conventional wisdom that a diagnosis of CP precluded the possibility of a genetic diagnosis [16], there are many individuals with CP who may benefit from genetic testing but have not received it.

We have previously defined subgroups of patients with CP who are likely to have a genetic diagnosis, therefore warranting genetic testing. Our team has categorized three different classifications for CP: “cryptogenic CP”, “non-cryptogenic CP”, and “CP masquerader” [17]. We defined CP masquerader as an individual with a diagnosis of CP, or with features suggestive of CP, but who has experienced developmental regression or progressive symptoms, which is incompatible with the definition of CP as a non-progressive motor encephalopathy. We defined non-cryptogenic CP as the diagnosis of CP in the presence of one or more known perinatal risk factors typically associated with CP (including prematurity, intraventricular/periventricular hemorrhage, hypoxic ischemic injury, and others, detailed in the Methods below) and absence of regression/progressive symptoms. We defined cryptogenic CP as the diagnosis of CP in the absence of any of these perinatal risk factors and the absence of regression/progressive symptoms. We have demonstrated that the diagnostic yield of genetic testing (in particular exome sequencing) is particularly high in the CP masquerader and cryptogenic CP categories [15,17] and developed a clinical approach for identifying patients with CP with these subtypes [15,17].

A major challenge is implementing into practice genetic testing for individuals most likely to have genetic causes of their CP. One issue may be clinicians who manage patients with CP (including pediatricians, neurologists, physiatrists, and orthopedics) in busy clinical settings have limited time during each visit to ascertain the presence/absence of all these aforementioned clinical features in order to classify patients into one of three classification groupings. Computational phenotyping may provide a potential informatics-based solution to this challenge. Computational phenotypes involve use of medical diagnostic billing codes from the electronic health record (EHR) to identify phenotypic features of a patient [18]. For example, our team has used computational phenotypes to help identify patients with features of the genetic neurodevelopmental disorder PTEN hamartoma tumor syndrome who may potentially benefit from genetic testing [19].

We hypothesized that computational phenotypes can classify patients with CP or CP-like conditions into etiological subgroups (CP masquerader, cryptogenic CP, and non-cryptogenic CP) with similar accuracy to manual chart review. To test this hypothesis, we developed a computational approach to defining CP masquerader, cryptogenic CP, and non-cryptogenic CP based on ICD-9/ICD-10 billing codes. We grouped the CP masquerader and cryptogenic CP categories (those who would benefit from genetic testing), distinct from the non-cryptogenic CP group that has low likelihood of a genetic diagnosis. We evaluated the predictive power of this algorithm to classify patients compared to manual chart review by applying it to a cohort of 250 individuals with cryptogenic/non-cryptogenic CP or CP masqueraders.

## Methods

### Participants

We used data from participants enrolled in the Boston Children’s Hospital (BCH) CP Sequencing Study, approved by the BCH Institutional Review Board, which performs next-generation sequencing on individuals with CP and CP masqueraders through the Children’s Rare Disease Cohorts (CRDC) initiative [20]. We selected this group for the present analysis, as many had undergone evaluations with multiple specialists in the Boston Children’s Hospital CP Clinic, ensuring a broad range of available billed ICD-9/ICD-10 codes.

Participants of any age or sex were eligible if they met standard criteria for CP, or if they could be considered to meet standard criteria for CP except for regression or progressive symptoms, which we had labeled as CP masquerader. Exclusion criteria included the presence of a molecular diagnosis which completely explained the clinical phenotype. Referrals to this study were from neurologists, orthopedists, physiatrists, nurse practitioners, complex care pediatricians, or neurosurgeons associated with the Boston Children’s Hospital CP Center who have extensive experience in management of CP.

Prospective elements of the study included recruitment, enrollment, sequencing, and categorization of participants as CP or CP masqueraders. Retrospective aspects of the study included classification of CP as cryptogenic/non-cryptogenic (see below under Overview of Classification Approach) and designation of primary CP motor phenotype (spastic hemiplegic, spastic diplegic, spastic quadriplegic, dyskinetic, or hypotonic-ataxic). These retrospective elements were based on review of the electronic medical record by a clinician board-certified in child neurology and neurodevelopmental disabilities (S.S.).

For this analysis, we selected the first 250 enrolled probands with an ascertained classification of CP (CP masquerader, cryptogenic CP, non-cryptogenic CP), an ascertained primary motor phenotype which fell into one of the 5 categories noted above, and available ICD-9/ICD-10 billing codes. We had previously characterized the CP phenotypes and research exome sequencing analysis results of the first 50 enrolled cases [17].

### Overview of Classification Approach

Our overall goal was to evaluate the performance of a computational approach using ICD-9/ICD-10 billing codes, compared to a manual review of the electronic medical record, in determining which patients with CP may warrant genetic testing (Figure 1). The test outcome was the classification of either cryptogenic CP or CP masquerader (as opposed to non-cryptogenic CP) in the participant.

**Figure 1.**
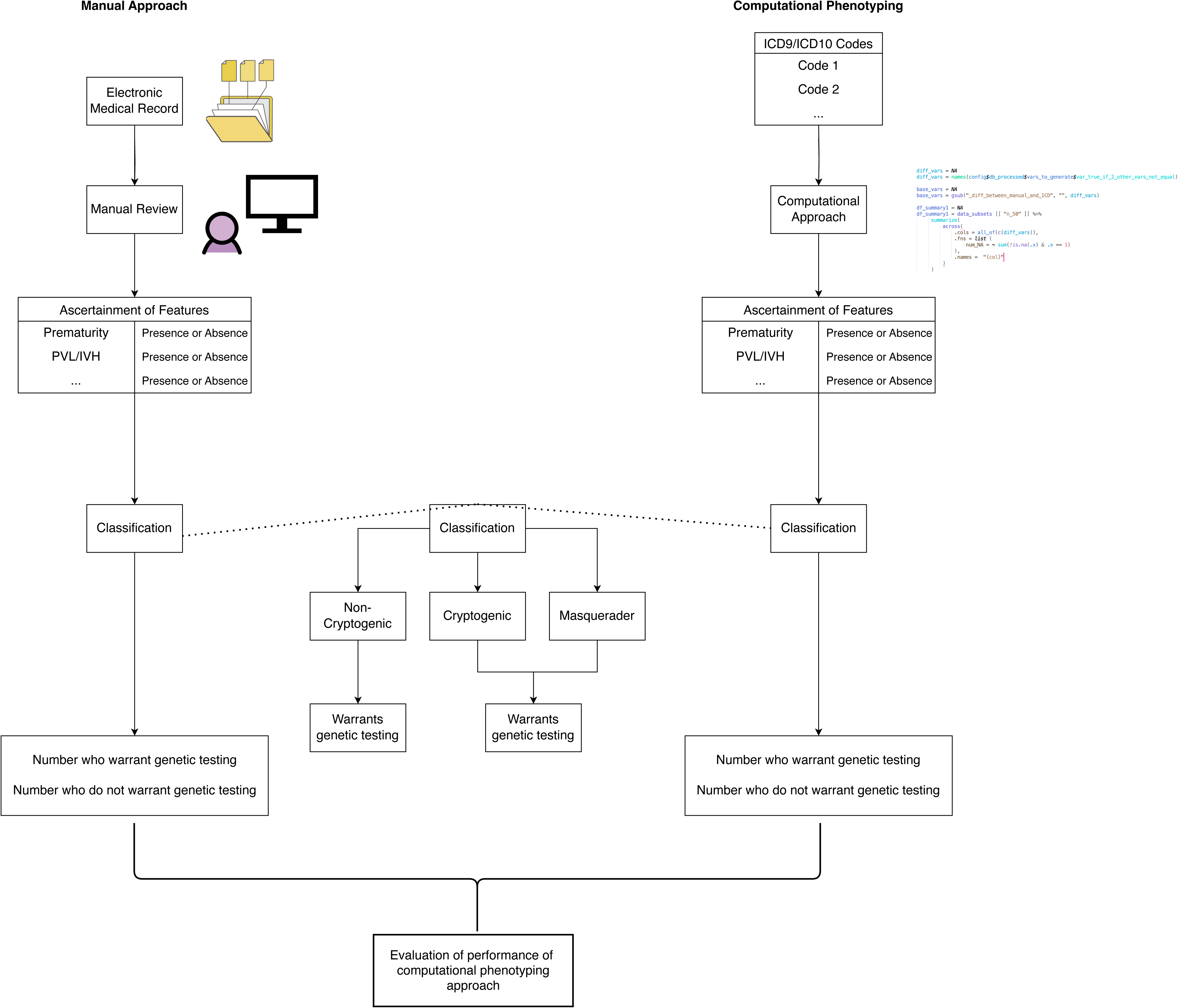
Schematic overview of process involving manual review of the electronic medical record as well as process involving computational phenotyping.

The classification of CP masquerader was based on the presence of progressive neurological symptoms (in the case of computational phenotyping) or the presence of developmental regression/progressive neurological symptoms (in the case of manual review). The reason for this difference (i.e., use of *both* regression and progressive neurological symptoms in the CP masquerader determination based on manual review but *only* progressive neurological symptoms in the CP masquerader determination based on computational phenotyping) is that while there is an ICD code for progressive neurological symptoms, there is not an appropriate ICD code for developmental regression. The ICD code R62.50 is sometimes used to denote regression, but its title is “unspecified lack of expected normal physiological development in childhood” which is not specific to developmental regression and could significant developmental delay.

The classification of non-cryptogenic CP was based on the following: (a) the absence of progressive neurological symptoms (in the case of computational phenotyping) or the absence of developmental regression/progressive neurological symptoms (in the case of manual review) (b) the presence of one or more of the following 15 clinical features that we have deemed *a priori* as CP risk factors:

1. prematurity (≤ 32 weeks gestation)
2. periventricular/intraventricular hemorrhage
3. intraparenchymal hemorrhage other than periventricular/intraventricular hemorrhage
4. perinatal stroke
5. evidence of other acute perinatal event (such as acute onset of decreased fetal movements)
6. hypoxic ischemic injury
7. neonatal encephalopathy other than due to hypoxic ischemic injury
8. kernicterus
9. fetal infection (i.e., TORCH infections)
10. maternal infection at delivery leading to sepsis in the mother
11. neonatal infection leading to sepsis
12. neonatal respiratory arrest
13. neonatal cardiac arrest
14. hydrocephalus (congenital or non-congenital)
15. traumatic brain injury

This list is a modification of the features we had presented previously [17]. Specifically, we had added neonatal encephalopathy other than that due to hypoxic ischemic injury.

The classification of cryptogenic CP was based on the following: (a) the absence of progressive neurological symptoms (in the case of computational phenotyping) or the absence of developmental regression/progressive neurological symptoms (in the case of manual review) and (b) the absence of any of these 15 clinical features.

### CP Classification Based on Manual Chart Review

For each participant, we manually reviewed medical documentation and explicitly denoted the presence/absence of the aforementioned 15 CP risk factors, as well as the presence/absence of developmental regression and progressive neurological symptoms (Figure 2). Based on review of presence/absence of these clinical features of interest, we designated participants with developmental regression or progressive neurological symptoms as CP masquerader. For patients who met standard criteria for CP, we further classified them into two categories: cryptogenic CP (no CP risk factors) and non-cryptogenic CP (one or more CP risk factors).

**Figure 2.**
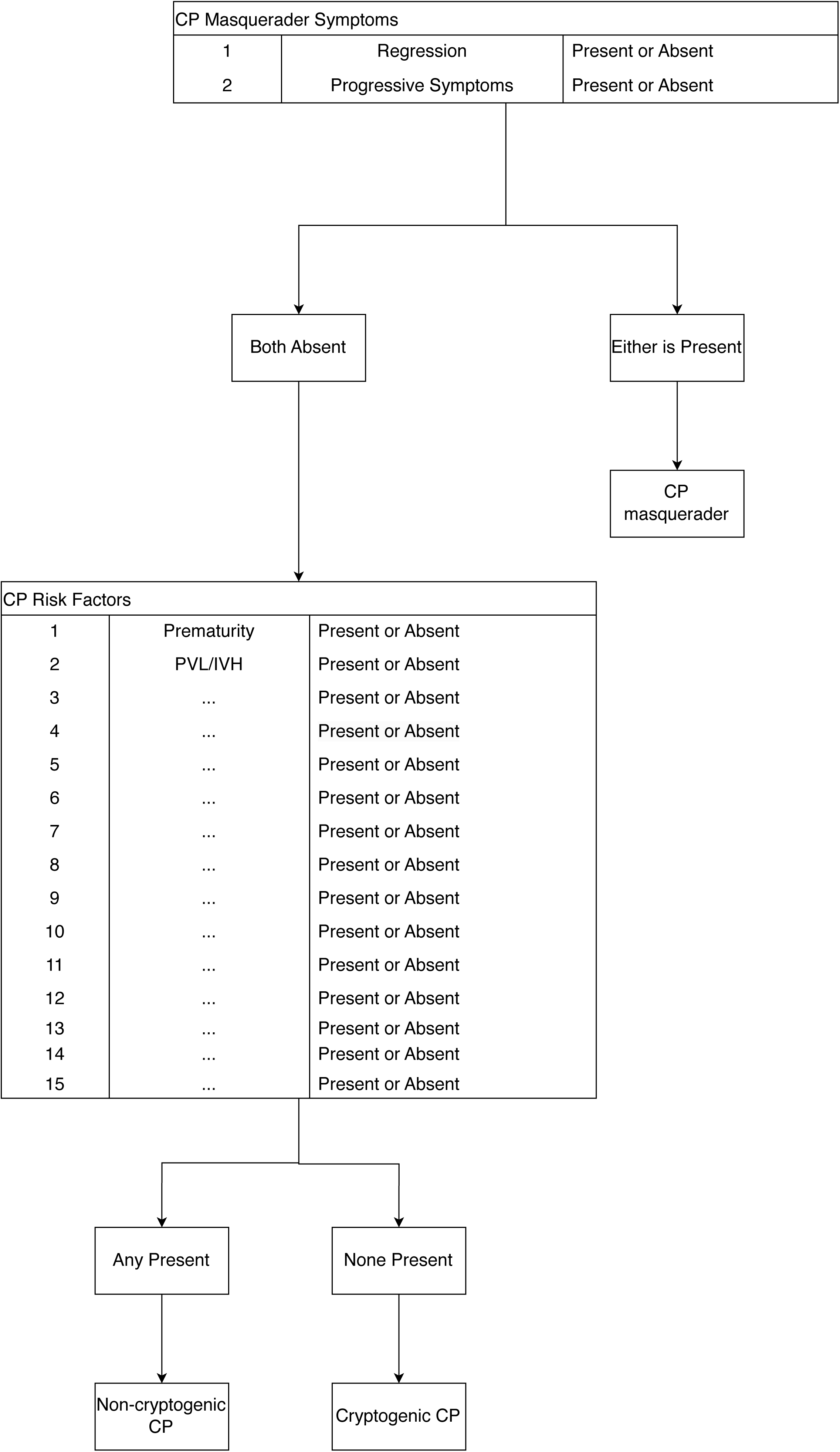
Details of how presence/absence of specific clinical features translate into the three specific CP classifications based on manual review of the electronic medical record.

### CP Classification Based on Computational Phenotype

We retrieved ICD-9 and ICD-10 codes for all participants using an automated extraction process applied to the electronical medical record. We created a dictionary mapping each clinical feature of interest (all 15 CP risk factors plus progressive symptoms) to a set of ICD-9/ICD-10 codes (Table S1). As noted previously, there was not a suitable ICD-9/ICD-10 code for developmental regression. Each ICD-9/ICD-10 billing code fell under precisely one clinical feature of interest.

We developed an algorithm for classifying CP etiology into one of the three categories using ICD- 9/ICD-10 billing codes (Figure S1). For each participant, the algorithm iterated through every billing code in the patient’s ICD-9/ICD-10 list, and if the billing code was present in the mapping dictionary, the algorithm marked the patient as having that specific clinical feature of interest.

Next, the algorithm determined if the participant had progressive symptoms based on ICD-9/ICD-10 codes review. If so, it classified the participant as CP masquerader based on ICD-9/ICD-10 codes review. Otherwise, it would determine if any of the CP risk factors were present based on ICD-9/ICD-10 codes review. If any of CP risk factors were present based on ICD-9/ICD-10 codes review, the participant received a classification of non-cryptogenic CP based on ICD-9/ICD-10 codes review, and if none of the CP risk factors was present based on ICD-9/ICD-10 codes review, the participant received a classification of cryptogenic CP based on ICD-9/ICD-10 codes review.

We implemented this algorithm in R (version 4.4.0).

### Comparison of CP Classification Based on Manual Review vs. Computational Phenotype

After we performed the processes outlined above, each participant had a CP classification (possible values: CP masquerader, non-cryptogenic CP, cryptogenic CP) based on manual review conducted by a board-certified child neurologist trained in neurodevelopmental disabilities (SS) and a CP classification based on computational phenotyping with ICD-9/ICD-10 codes. Since individuals who are classified as CP masquerader or cryptogenic CP both require genetic testing, and for ease of comparison, we regrouped the three classes (CP masquerader, non-cryptogenic CP, cryptogenic CP) into two classes: genetic testing warranted (CP masquerader or cryptogenic CP) and genetic testing not warranted (non-cryptogenic CP). We evaluated the performance of the computational phenotyping approach with sensitivity, specificity, positive predictive value, and negative predictive value. We designated the classification based on manual review as the truth set and classification based on computational phenotyping with ICD-9/ICD-10 codes as the prediction set. Moreover, the classification of “genetic testing warranted” represented the positive class.

## Results

In our cohort of 250 probands, there were 143 males and 107 females. Average age of enrollment of participants was 9.2 years (standard deviation = 6.5 years). Of note, this cohort of 250 individuals included data from 49/50 of the participants who our team had previously characterized from the standpoint of sequencing analysis [17]. There was one participant from that initial cohort of 50 who we had excluded from the present analysis due to limited perinatal history leading to inability to delineate CP classification. Moreover, we had updated the CP classification for two previously reported participants from cryptogenic CP to CP masquerader based on new information regarding regression and/or progressive symptoms.

Based on manual chart review, 8% (19/250) had CP masquerader classification, 42% (105/250) had cryptogenic CP classification, and 50% (126/250) had non-cryptogenic CP classification. Of the 19 participants with CP masquerader classification, all but 6 had progressive neurological symptoms on manual review; the other 6 participants had regression but not progressive neurological symptoms on manual review. In addition, among the 19 participants with CP masquerader classification, there were 6 who each had one of the 14 CP risk factors on manual review (n = 3 with prematurity, n = 1 with hypoxic ischemic injury, n = 1 with perinatal stroke, n = 1 with fetal infection).

Among the 125 participants with non-cryptogenic CP, the mean number of present CP risk factor features based on manual review was 1.5 (sd = 0.8, minimum = 1, maximum = 5). Each of the 15 CP risk factor features, except for fetal infection and traumatic brain injury, had representation by at least one participant with non-cryptogenic CP. Of note, fetal infection based on manual review was noted in one participant in the CP masquerader classification; traumatic brain injury based on manual review was noted in none of the participants in the cohort.

Based on computational phenotyping with ICD-9/ICD-10 codes, 9% (22/250) had CP masquerader, 52% (131/250) had cryptogenic CP classification, and 38% (97/250) had non-cryptogenic CP classification.

We evaluated the performance of the computational phenotyping approach in accurately distinguishing between the two genetic testing classification groups, genetic testing warranted (CP masquerader or cryptogenic CP) and genetic testing not warranted (non-cryptogenic CP). The computational phenotyping process had a sensitivity of 95%, specificity of 72%, a positive predictive value of 77%, and a negative predictive value of 94%.

In addition, for the entire cohort, for each feature (CP risk factor or progressive symptoms), we calculated the sensitivity (number individuals who had the feature based on both manual review and ICD- 9/ICD-10 codes review, divided by the number of individuals who had the feature based on manual review) (Figure 3). The features associated with the top 5 highest sensitivities were: fetal infection (100%, 1/1), hypoxic ischemic injury (23/26), hydrocephalus (12/14), intraparenchymal hemorrhage other than periventricular/intraventricular hemorrhage (5/6), and prematurity (52/71). The features associated with sensitivities of 0% were: perinatal respiratory arrest, perinatal cardiac arrest, and maternal infection leading to sepsis.

**Figure 3.**
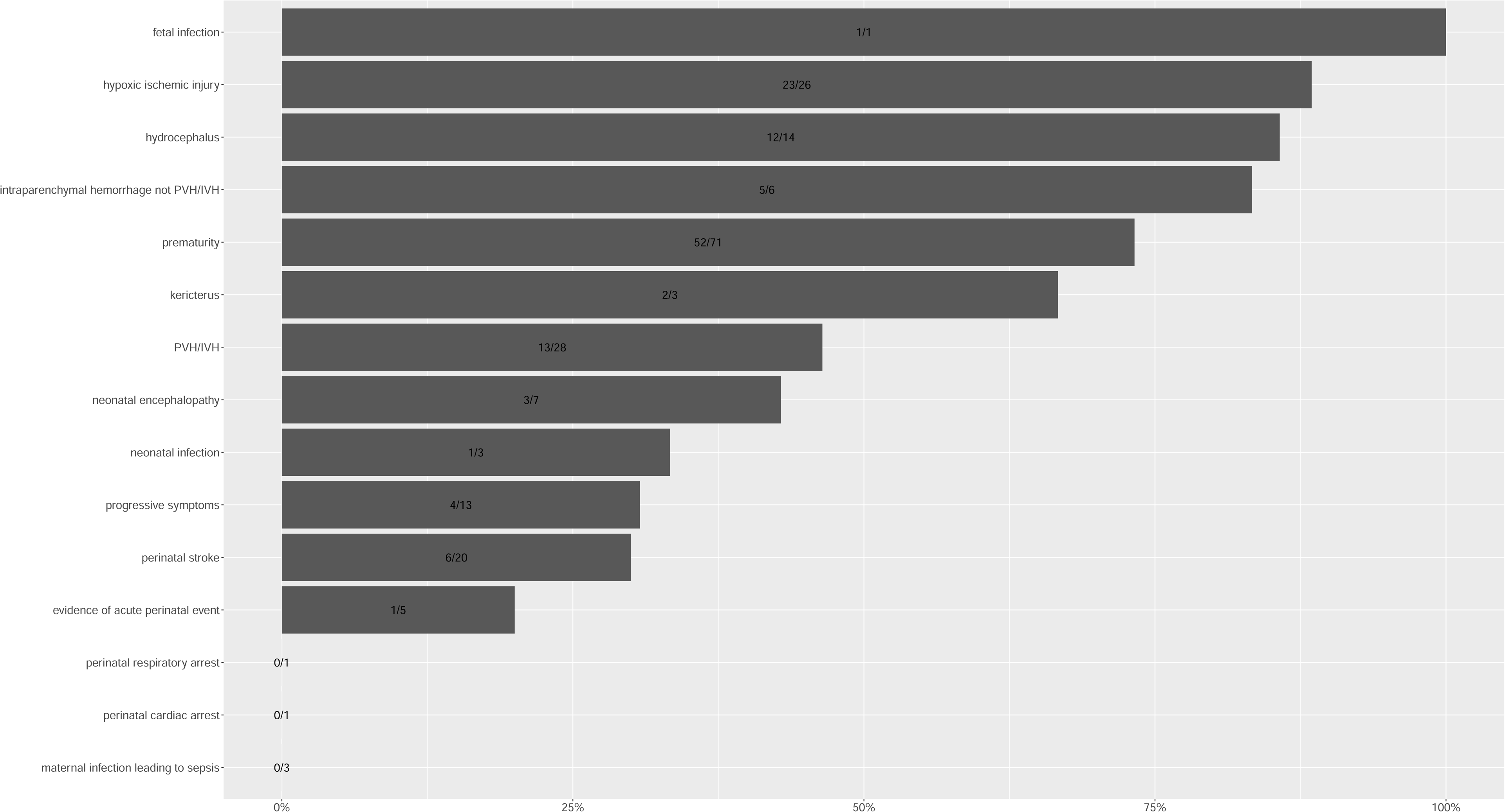
Sensitivity of the computational approach for detecting each of the 14 CP risk factors and progressive symptoms. In this context, sensitivity is the number individuals who had the feature based on both manual review and ICD-9/ICD-10 codes review, divided by the number of individuals who had the feature based on manual review.

## Discussion

We have demonstrated the feasibility of utilizing computational phenotyping for determining subgroups of CP and related disorders who may warrant genetic testing. Moreover, we have shown that this approach has a high positive predictive value, i.e., the likelihood that a person with a positive classification of warranting genetic testing by ICD-9/ICD-10 codes actually falling into the cryptogenic CP/CP masquerader classification.

This computational approach to identifying subsets of CP and CP-related conditions may serve as a clinical decision support (CDS) tool for use in electronic medical record systems [21]. For example, if a clinician is seeing a patient with CP and is using an electronic medical record system, there could be a user-facing alert triggered if the patient has cryptogenic CP or a CP masquerading condition based on prior billing codes but has not had genetic testing. This alert could prompt the user to review the medical history to ascertain whether genetic testing may be warranted.

A CDS tool utilizing ICD billing codes offers advantages over routine clinical practice in certain scenarios. The first is as follows. We acknowledge that the gold standard of ascertaining CP etiological classification is through careful review of the medical history, complete physical examination, and review of neuroimaging and other diagnostics. However, in a busy clinical practice, especially if a patient has an existing diagnosis of CP made by a different provider, such attention may be missed. This tool could serve as a prompt for consideration of etiology. Secondly, a computational tool based on billing codes embedded in an electronic medical record system can run in real-time using longitudinal data and does not depend on any single provider’s billing data. For instance, a patient with non-cryptogenic CP due to periventricular leukomalacia and prematurity may develop progressive worsening neurological symptoms over time, thus fitting into the CP masquerader classification. If one clinician documents and bills this feature, yet another clinician seeing the patient at a later timepoint does not realize progressive neurological symptoms have occurred, a computational phenotype approach-based CDS tool would potentially prompt consideration of genetic testing for CP masquerader status. Third, each subspecialist seen by a patient with CP may focus on pertinent features relevant to that subspecialist and may miss other aspects of the presentation, including whether genetic testing is warranted. For example, if a patient with CP and epilepsy sees an epileptologist, the focus during that visit would be appropriately on seizure management and less so on CP risk factors. An alert in the electronic medical record system based on billing codes can remind that subspecialist to consider genetic testing if warranted.

Establishing a genetic diagnosis in a person with CP or related disorders is crucial for many aspects of clinical management. The genetic landscape of CP and CP-masquerading conditions includes a growing number of Mendelian and chromosomal disorders. Data from a systematic review and Delphi process support the notion there is actionability that may result from genetic findings in CP [22]. Our group has shown in patients with CP and “CP masquerading” conditions found to have an underlying genetic diagnosis, a substantial percentage undergo etiological diagnosis-influenced management changes, including medication interventions, surveillance impact, family member testing, and patient education [23]. In other words, consideration of genetic testing in persons with CP in certain circumstances is essential, and novel approaches which can enhance utilization of genetic testing are much needed.

Although computational phenotyping offers several advantages, there are notable limitations. One such issue is that while it is a computational-based algorithm that does reduce human error, it still relies on ICD-9/10 billing codes to be input manually by clinicians. These ICD-9/10 billing codes may be inaccurate due to human error, or due to the fact that these billing codes may sometimes only be able to approximate certain features.

Limitations aside, a computational approach may be applied to larger datasets spanning an entire hospital or even multiple hospital systems, helping identify patients with CP who warrant genetic testing. This will make more patients potentially eligible for gene-based treatments as they emerge in addition to providing diagnostic precision, closure for families, and a multitude of other benefits.

## Data Availability

Data is available upon request to the corresponding author.

## Data Statement

Data is available upon request to the corresponding author.

## Conflicts of Interest

**SS:** SS has received grants from NIH/NINDS (K23NS119666); consulting fees from GLG, Guidepoint (which connected to a client, Fortress Biotech), Novartis, ExpertConnect, Orchard Therapeutics.

---

**Figure S1.**
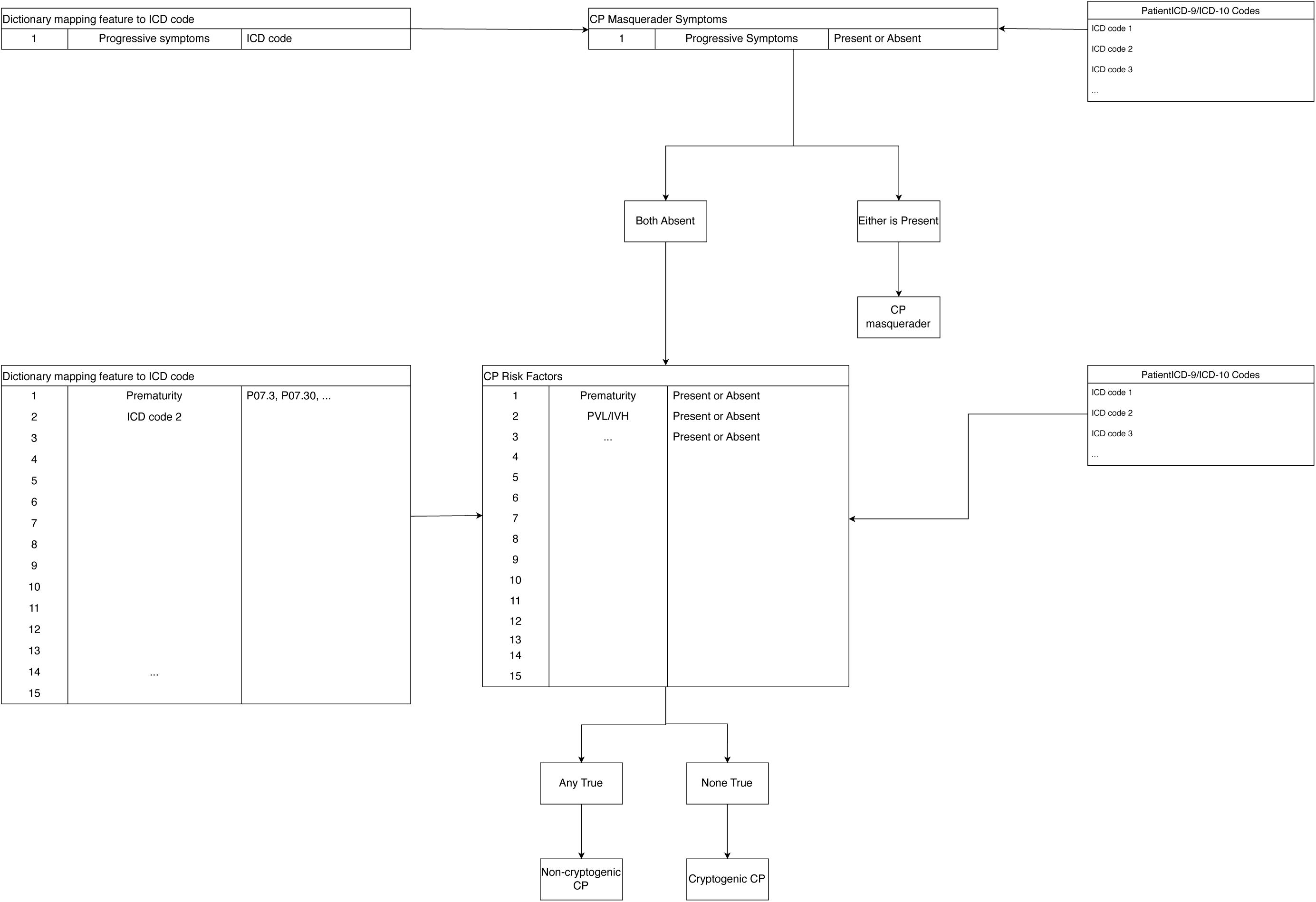
Details of how presence/absence of specific ICD-9/ICD-10 codes translate into the three specific CP classifications based on computational phenotyping.

**Table S1.**
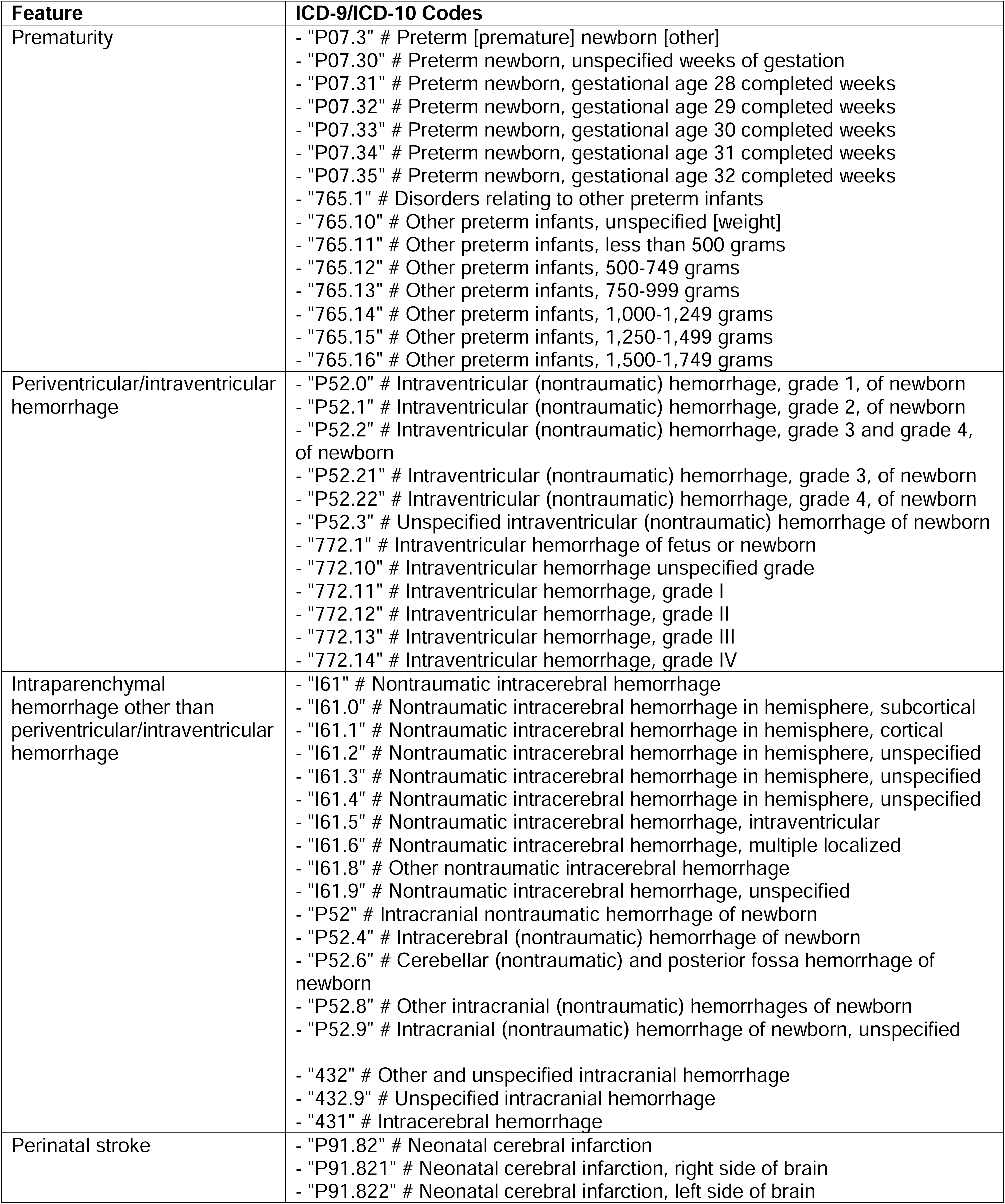
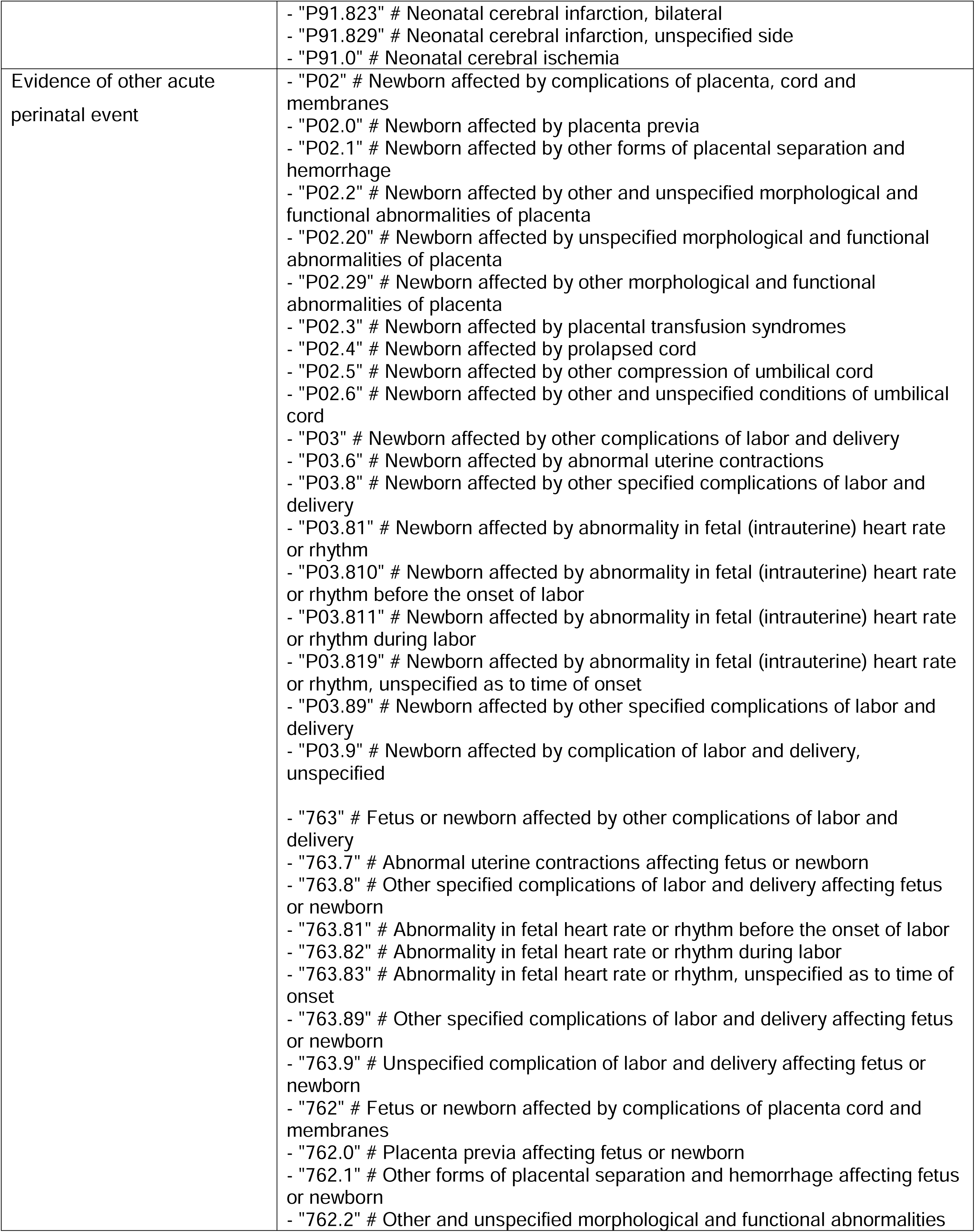
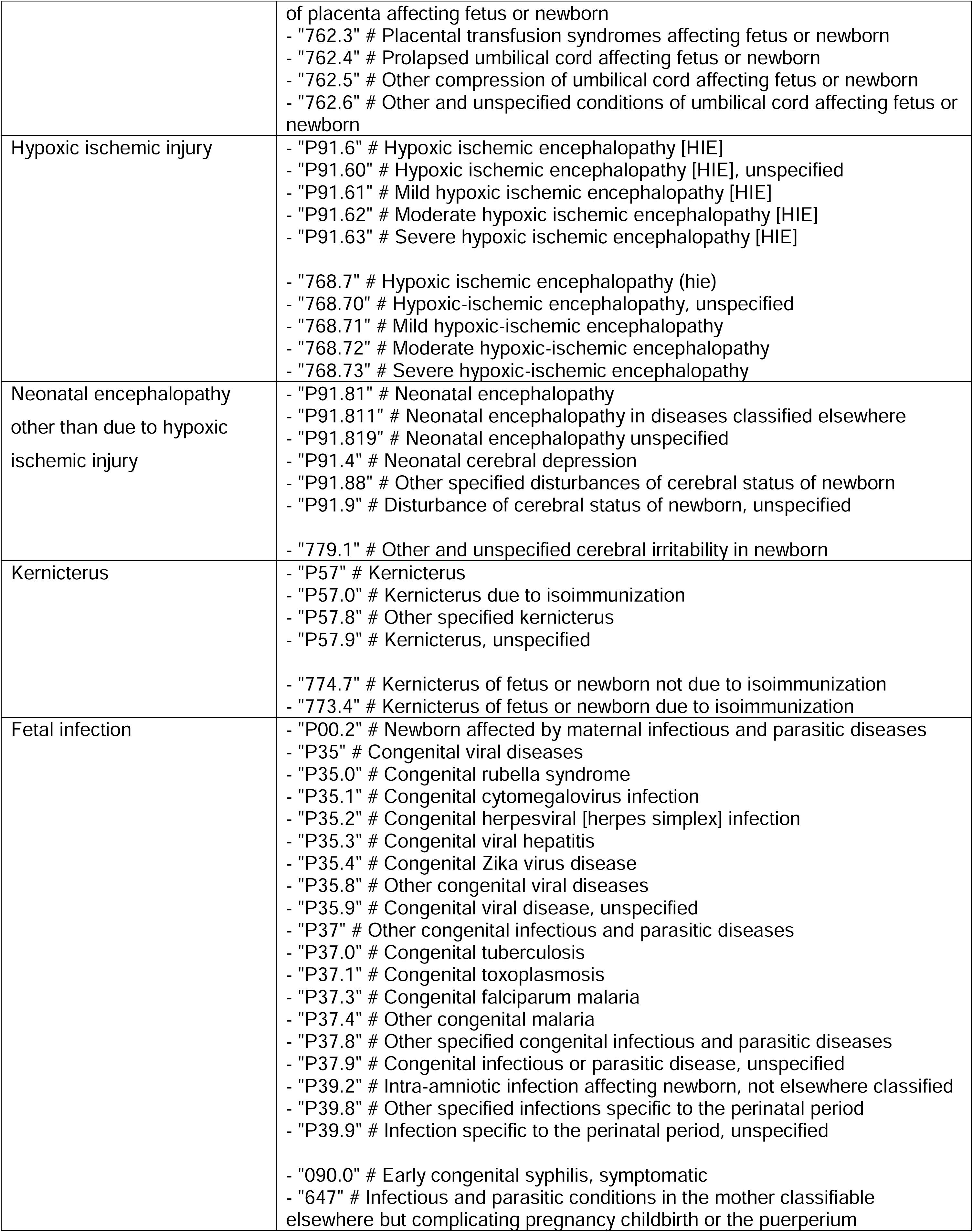
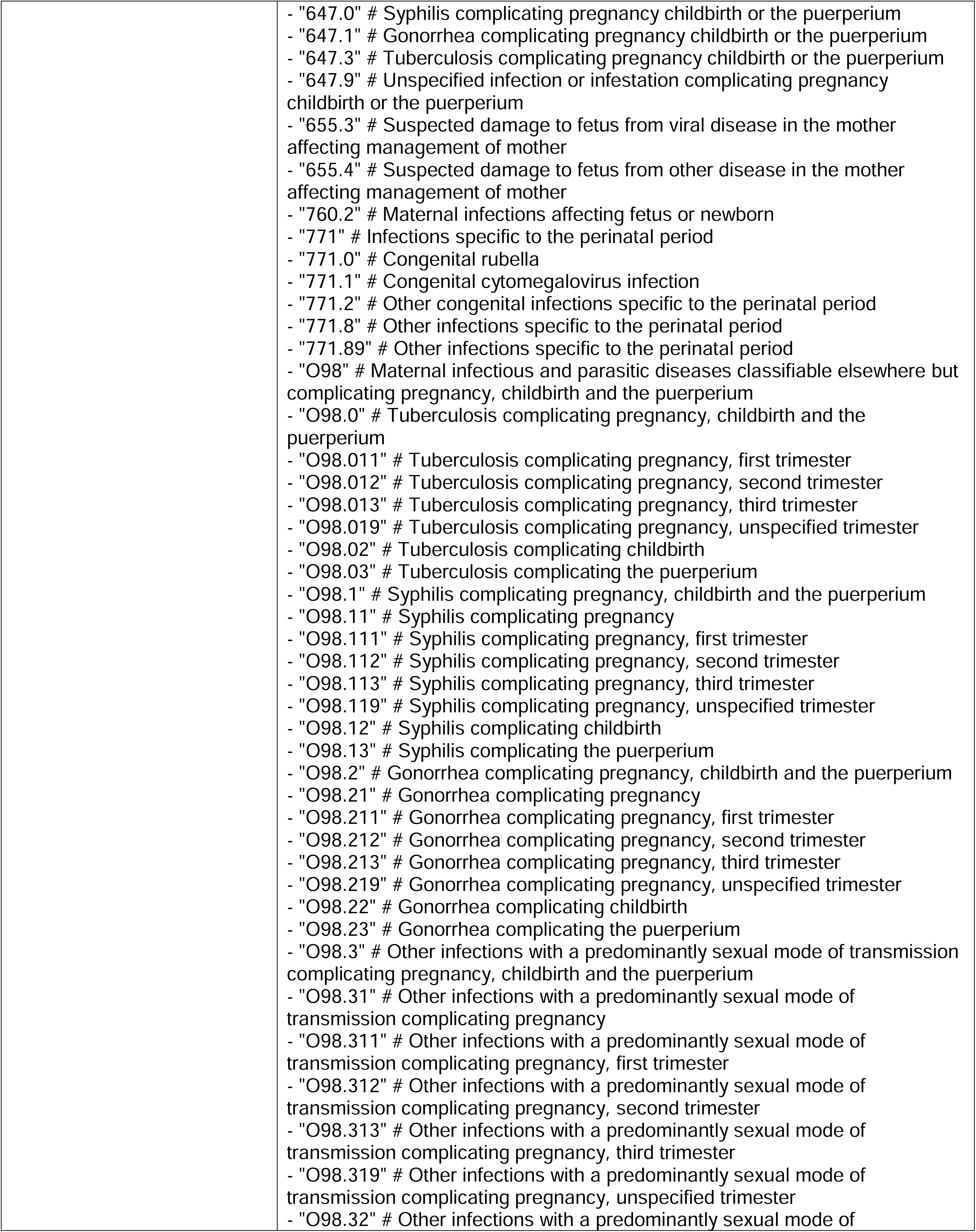
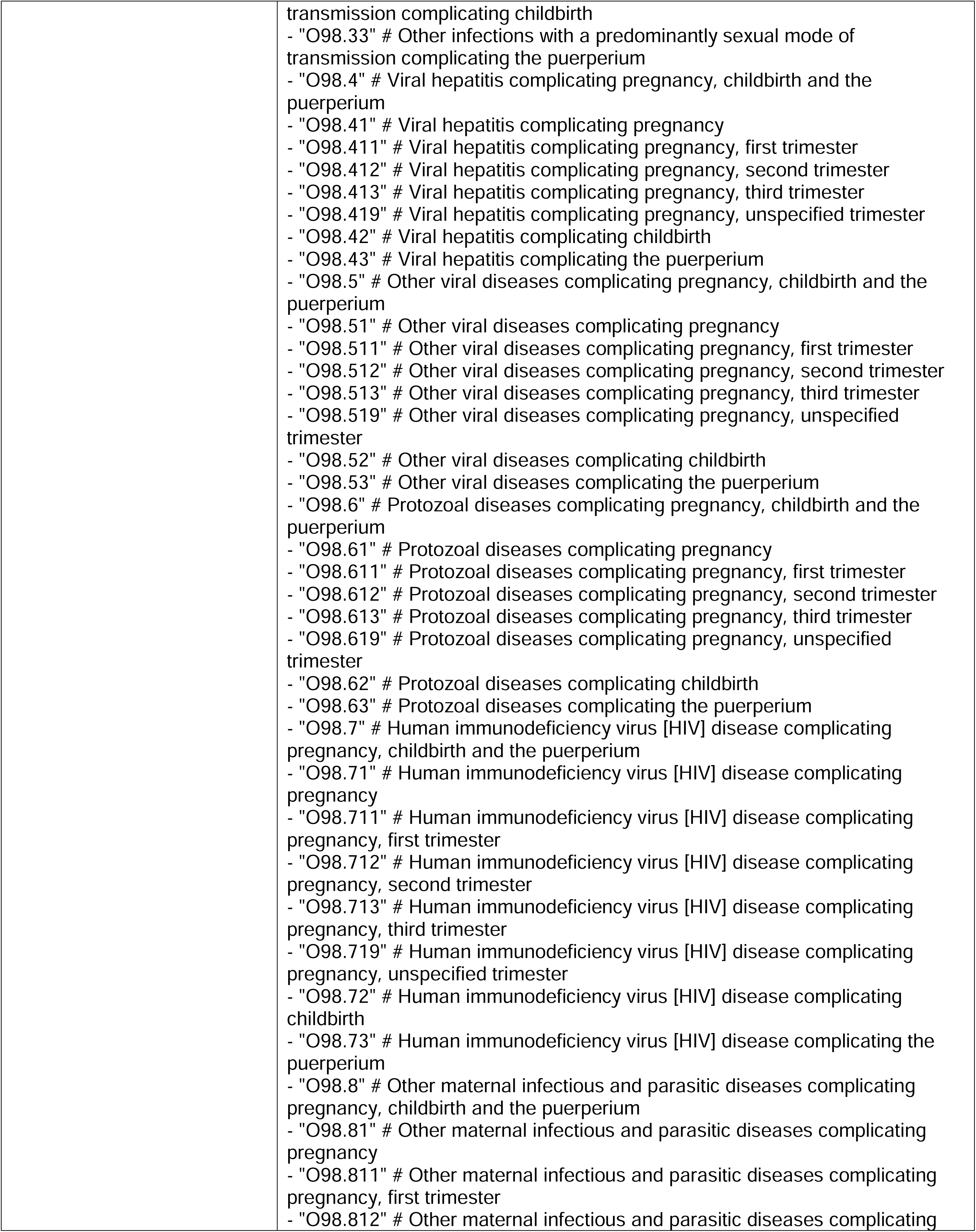
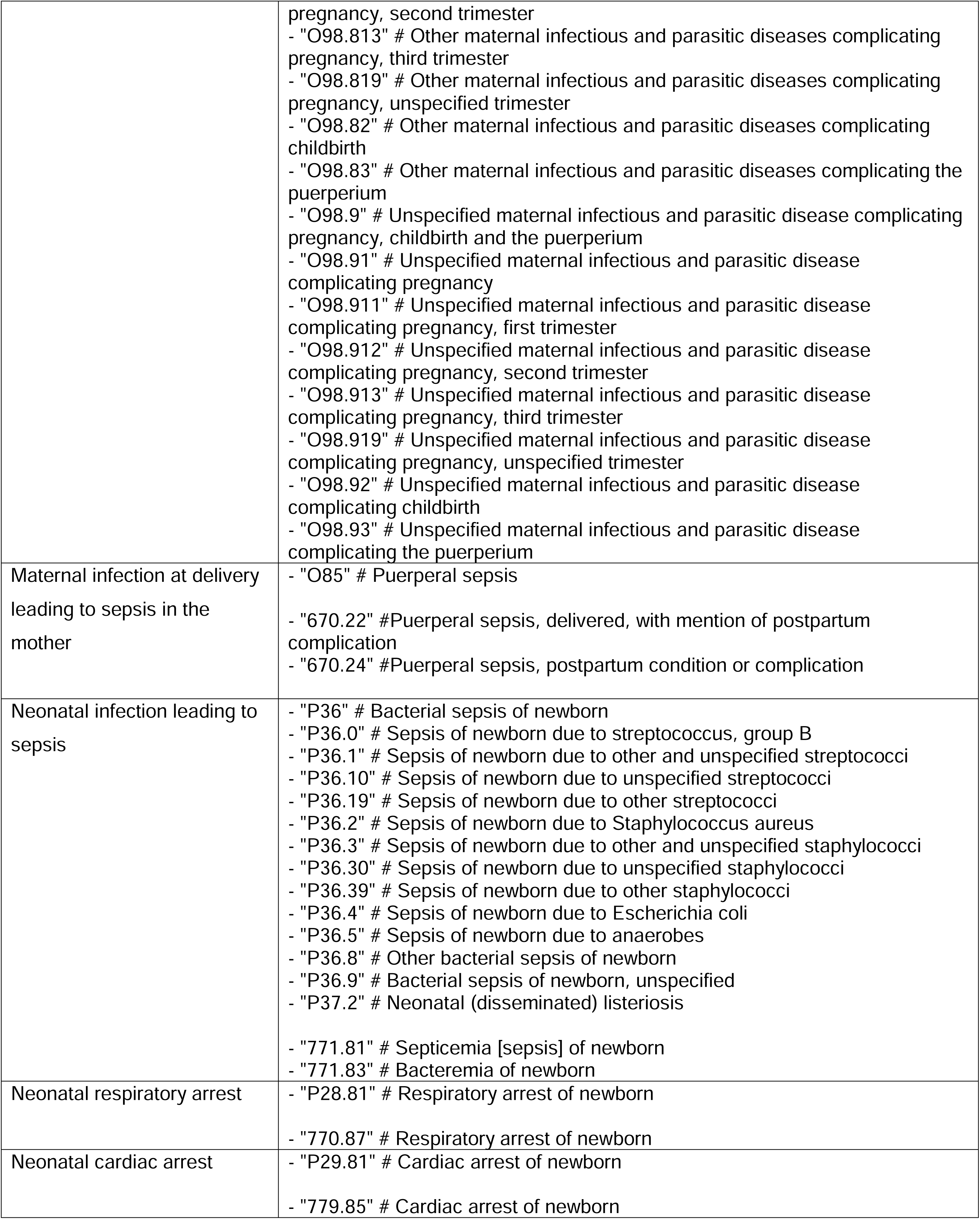
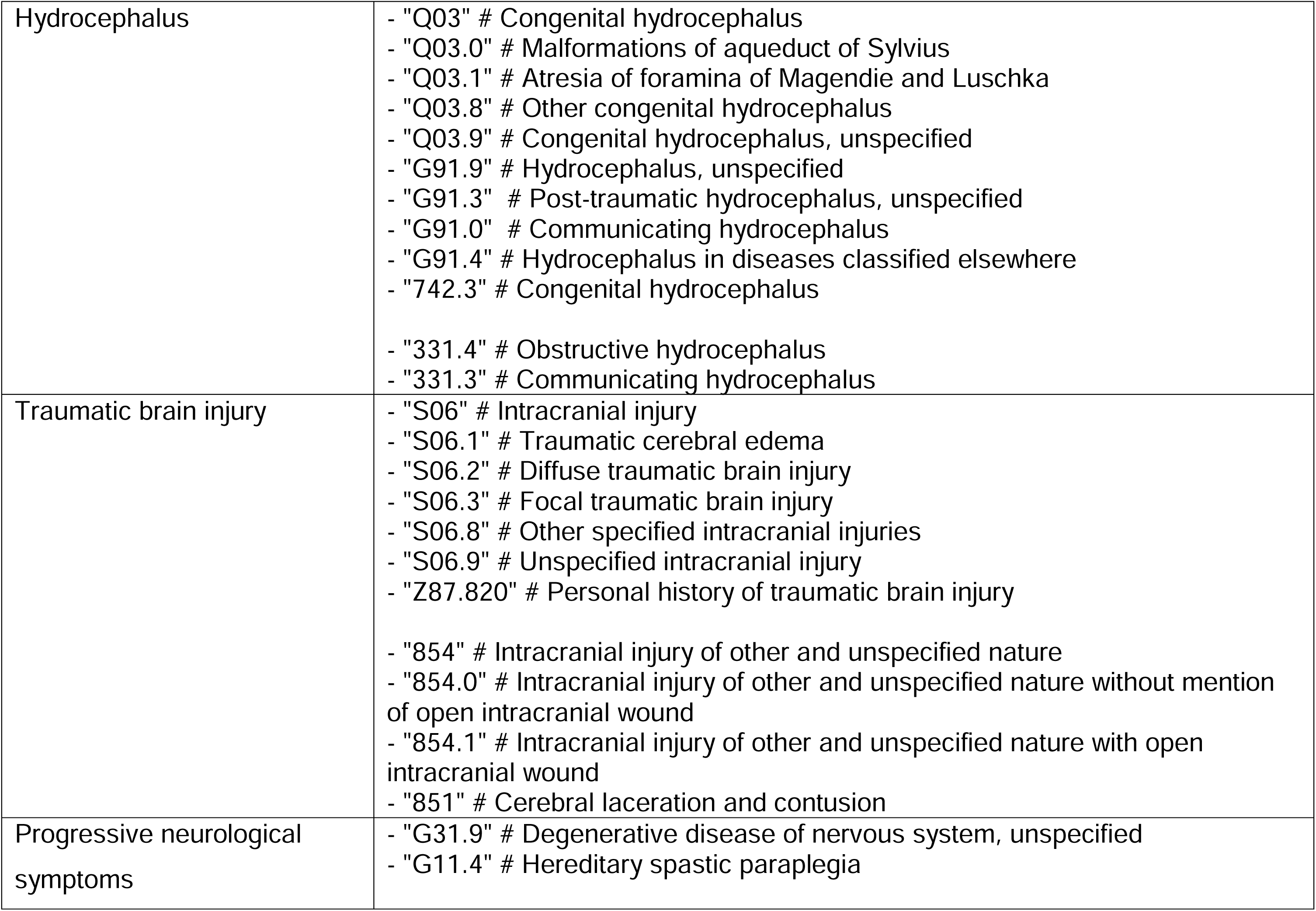
Mapping of ICD-9/ICD-10 codes to different clinical features.

## Notes

### Author Declarations

We used data from participants enrolled in the Boston Children's Hospital (BCH) CP Sequencing Study, approved by the BCH Institutional Review Board, which performs next-generation sequencing on individuals with CP and CP masqueraders through the Children's Rare Disease Cohorts (CRDC) initiative.

